# Meta-analysis of diagnostic performance of serological tests for SARS-CoV-2 antibodies and public health implications

**DOI:** 10.1101/2020.05.03.20084160

**Authors:** Saverio Caini, Federica Bellerba, Federica Corso, Angélica Díaz-Basabe, Gioacchino Natoli, John Paget, Federica Facciotti, Sara Raimondi, Domenico Palli, Luca Mazzarella, Pier Giuseppe Pelicci, Paolo Vineis, Sara Gandini

## Abstract

Serology-based tests have become a key public health element in the COVID-19 pandemic to assess the degree of herd immunity that has been achieved in the population. These tests differ between one another in several ways. Here, we conducted a systematic review and meta-analysis of the diagnostic accuracy of currently available SARS-CoV-2 serological tests, and assessed their real-world performance under scenarios of varying proportion of infected individuals. We included independent studies that specified the antigen used for antibody detection and used quantitative methods. We identified nine independent studies, of which six were based on commercial ELISA or CMIA/CLIA assays, and three on in-house tests. Test sensitivity ranged from 68% to 93% for IgM, from 65% to 100% for IgG, and from 83% to 98% for total antibodies. Random-effects models yielded a summary sensitivity of 82% (95%CI 75–88%) for IgM, and 85% for both IgG (95%CI 73–93%) and total antibodies (95%CI 74–94%). Specificity was very high for most tests, and its pooled estimate was 98% (95%CI 92–100%) for IgM and 99% (95%CI 98–100%) for both IgG and total antibodies. The heterogeneity of sensitivity and specificity across tests was generally high (I^2^>50%). In populations with a low prevalence (≤5%) of seroconverted individuals, the positive predictive value would be ≤88% for most assays, except those reporting perfect specificity. Our data suggest that the use of serological tests for large-scale prevalence surveys (or to grant “immunity passports”) are currently only justified in hard-hit regions, while they should be used with caution elsewhere.

## Introduction

Testing of patients for ongoing infection with SARS-CoV-2 is mostly conducted by detecting viral RNA in airways specimens using RT-PCR-based tests. These tests may prove less helpful in quantifying the actual number of COVID-19 cases in the population, as a large proportion of infected individuals are thought to be asymptomatic [1–2] or may not seek medical care because of mild symptoms, thus going unnoticed by surveillance systems and public health entities. Moreover, once the infection is resolved, RT-PCR tests are not informative for the previous infection. In order to overcome these shortcomings, serology-based tests are being increasingly used with the aim of gaining greater detail into the true prevalence of COVID-19 and to assess the degree of herd immunity that has been achieved in the population. Serology-based tests have thus become a key public health element in the COVID-19 pandemic and there has been a rapid growth in the number of available SARS-CoV-2 serological tests over recent months. These tests differ between one another in several ways, including the antigens used for antibody detection, the type of antibodies identified, and the laboratory method. Here, we conducted a systematic review and meta-analysis of the diagnostic accuracy of currently available SARS-CoV-2 serological tests, and assessed what their real-world performance under scenarios of varying proportion of infected individuals.

## Methods

We carried out a systematic literature search (updated to April 19^th^) to review scientific articles and technical manuals (referred to as “studies” henceforth) on immunological tests for detection of SARS-CoV-2 antibodies. We considered independent studies that specified the antigen used for antibody detection, used quantitative methods, and reported the number of true positives, true negatives, false positives, and false negatives. This information was extracted from each study alongside with the laboratory method used as reference. From studies reporting results for two different kits, we entered data for the “Beijing Wantai” kit (instead of the “Xiamen InnoDx Biotech” kit), for consistency with other studies. When two different antigens were tested, we entered data for the nucleocapsid (N) protein instead of the Spike protein, because they generally showed better sensitivity. Sensitivity analyses were conducted to assess the robustness of pooled results against these choices.

Based on the 2×2 contingency table, we calculated the test sensitivity and specificity (with 95% confidence intervals [CI]) and the diagnostic odds ratio (DOR), to provide an overall measure of the test performance [3]. We then calculated the positive (PPV) and negative (NPV) predictive values assuming a true prevalence of 5%, 10% and 20%. Pooled estimates of sensitivity and specificity were obtained through random-effects models after Freeman-Tukey double arcsine transformation. DOR were pooled by fitting a bivariate model which takes into account the correlation between sensitivity and specificity and uses their log-transformed values as normally distributed variables. Between-studies heterogeneity was assessed using the I^2^ statistics, which quantifies the percentage of variation attributable to heterogeneity rather than chance. An I^2^ below 50% was considered as an indicator of acceptable heterogeneity.

## Results

We identified nine studies [4–12] (Figure 1), of which six were based on commercial ELISA or CMIA/CLIA assays, and three on in-house tests, for detecting SARS-CoV-2 antibodies (Table 1). Most studies evaluated sensitivity and specificity separately for IgG and IgM, while only some reported those values for total antibodies. Only one study tested IgA (Euroimmun kit) [10]. Real-time RT-PCR was always used as the reference method for sensitivity, while the definition of negative subjects varied across studies (eTable 1 in the Supplement).

**Figure 1.**
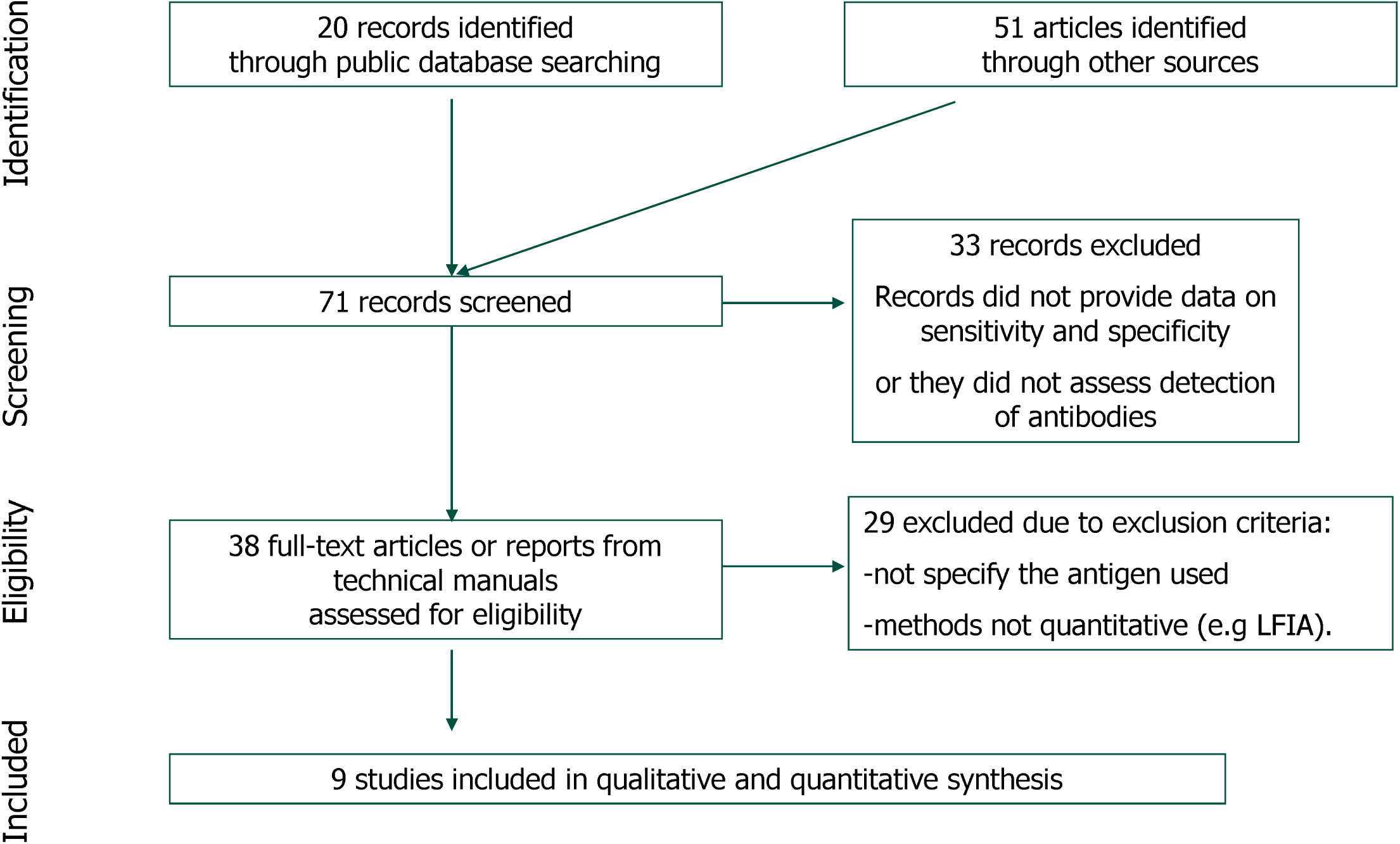
Flow-chart of the literature search of the diagnostic performance of serological tests for SARS-CoV-2 antibodies.

**Table 1.** Main characteristics of studies included in the systematic reviews and meta-analysis of the performance of currently available serological tests for SARS-CoV-2, along with test sensitivity and specificity and positive (PPV) and negative (NPV) predictive values assuming a true prevalence of 5%, 10% and 20%.

| Study/Report | Tests kit | Antibodies detected | No. of subjects | Sens | Spec | PPV-5 | PPV-10 | PPV-20 | NPV-5 | NPV-10 | NPV-20 | Test used | Antigen for detection |
| --- | --- | --- | --- | --- | --- | --- | --- | --- | --- | --- | --- | --- | --- |
| Lou et al. [4] | Beijing Wantai | Total Ab | 380 | 0.98 | 1.00 | 1.00 | 1.00 | 1.00 | 1.00 | 1.00 | 0.99 | ELISA | RBD domain |
|  | Xiamen InnoDx Biotech | Total Ab | 380 | 0.96 | 0.99 | 0.88 | 0.94 | 0.97 | 1.00 | 1.00 | 0.99 | CMIA | RBD domain |
|  | Beijing Wantai | IgM | 380 | 0.93 | 1.00 | 1.00 | 1.00 | 1.00 | 1.00 | 0.99 | 0.98 | ELISA | RBD domain |
|  | Xiamen InnoDx Biotech | IgM | 380 | 0.86 | 0.99 | 0.87 | 0.93 | 0.97 | 0.99 | 0.98 | 0.97 | CMIA | RBD domain |
|  | Beijing Wantai | IgG | 180 | 0.89 | 1.00 | 1.00 | 1.00 | 1.00 | 0.99 | 0.99 | 0.97 | ELISA | N protein |
| Lin et al. [5] |  | IgG | 159 | 0.82 | 0.98 | 0.63 | 0.79 | 0.89 | 0.99 | 0.98 | 0.96 | CLIA | N protein |
|  |  | IgM | 159 | 0.82 | 0.81 | 0.19 | 0.33 | 0.52 | 0.99 | 0.98 | 0.95 | CLIA | N protein |
| Liu et al. [6] |  | IgG | 314 | 0.74 | 1.00 | 1.00 | 1.00 | 1.00 | 0.99 | 0.97 | 0.94 | ELISA | Spike protein |
|  |  | IgM | 314 | 0.77 | 1.00 | 1.00 | 1.00 | 1.00 | 0.99 | 0.98 | 0.95 | ELISA | Spike protein |
|  |  | IgG | 314 | 0.70 | 1.00 | 1.00 | 1.00 | 1.00 | 0.98 | 0.97 | 0.93 | ELISA | N protein |
|  |  | IgM | 314 | 0.68 | 1.00 | 1.00 | 1.00 | 1.00 | 0.98 | 0.97 | 0.93 | ELISA | N protein |
| Zhao et al. [7] | Beijing Wantai | Total Ab | 386 | 0.93 | 0.99 | 0.84 | 0.92 | 0.96 | 1.00 | 0.99 | 0.98 | ELISA | RBD domain |
|  |  | IgG | 370 | 0.65 | 0.99 | 0.77 | 0.88 | 0.94 | 0.98 | 0.96 | 0.92 | ELISA | N protein |
|  |  | IgM | 386 | 0.83 | 0.99 | 0.76 | 0.87 | 0.94 | 0.99 | 0.98 | 0.96 | ELISA | RBD domain |
| Creative Diagnostics [8] |  | IgG | 46 | 1.00 | 1.00 | 1.00 | 1.00 | 1.00 | 1.00 | 1.00 | 1.00 | ELISA | Whole virus lysate |
| Epitope Diagnostic [9] |  | IgG | 84 | 1.00 | 1.00 | 1.00 | 1.00 | 1.00 | 1.00 | 1.00 | 1.00 | ELISA | N protein |
| Lassaunière et al. [10] | Beijing Wantai | Total Ab | 112 | 0.93 | 1.00 | 1.00 | 1.00 | 1.00 | 1.00 | 0.99 | 0.98 | ELISA | RBD domain |
|  | Euroimmun | IgA | 112 | 0.93 | 0.93 | 0.41 | 0.60 | 0.77 | 1.00 | 0.99 | 0.98 | ELISA | RBD domain |
|  |  | IgG | 112 | 0.67 | 0.96 | 0.47 | 0.65 | 0.81 | 0.98 | 0.96 | 0.92 | ELISA | RBD domain |
| Ortho-Clinical Diagnostics [11] |  | Total Ab | 436 | 0.83 | 1.00 | 1.00 | 1.00 | 1.00 | 0.99 | 0.98 | 0.96 | ELISA | Spike protein |
| Adams et al. [12] |  | IgG | 90 | 0.85 | 1.00 | 1.00 | 1.00 | 1.00 | 0.99 | 0.98 | 0.96 | ELISA | Spike protein |
|  |  | IgM | 90 | 0.70 | 1.00 | 1.00 | 1.00 | 1.00 | 0.98 | 0.97 | 0.93 | ELISA | Spike protein |

Sens: Sensitivity. Spec: specificity. PPV-5, PPV-10, and PPV-20: Positive Predictive Values at a hypothesized prevalence of 5%, 10% and 20%. NPV-5, NPV-10, and NPV-20:
Negative Predictive Values at a hypothesized prevalence of 5%, 10% and 20%.
ELISA: enzyme-linked immunosorbent assay. CMIA: chemiluminescent microparticle immunoassay. CLIA: chemiluminescent immunoassay.

The reviewed studies had sample sizes ranging between 46 and 436 subjects. For IgM, sensitivity ranged from 68% in Liu et al. (in-house test) to 93% in Lou et al. (Beijing Wantai kit), based on 314 and 380 subjects, respectively. The lowest sensitivity for IgG detection (65%) was in Zhao et al. (Bejiing Wantai kit), which tested 370 subjects, while two smaller-size studies (46 and 84 subjects) reached a sensitivity of 100% [8–9].

For IgM testing, the PPV had lowest values of 19% to 52% (in the 5% and 20% true-prevalence scenarios, respectively) in the study by Lin et al. (n=159), while it was 100% in all scenarios in the larger study (n=314) by Liu et al. For IgG, the PPV ranged between 47% and 81% (depending on the assumed prevalence) in the study by Lassaunière et al. (n=112), while it achieved 100% in Lou et al. (n=380). The NPV fell in the range 96–100% for all IgG and IgM kits when the prevalence was assumed to be 10% (the lower limit of the range became 98% and 92% for the 5% and 20% true-prevalence scenarios, respectively).

Meta-analysis yielded a summary sensitivity of 82% (95%CI 75–88%) for IgM, and 85% for both IgG (95%CI 73–93%) and total antibodies (95%CI 74–94%) (Table 2 and eFigures 1–6 in the Supplement). Pooled specificity was 98% (95%CI 92–100%) for IgM and 99% (95%CI 98–100%) for both IgG and total antibodies. Heterogeneity was consistently high (I^2^>50%), except when pooling specificity of IgG tests (I^2^=13%). Mostly due to the low proportion of false positives, the pooled DOR was generally very high (≈2,800 for IgM, and ≈1,300 for IgG and total antibodies). Results remained substantially unaltered in all sensitivity analyses (data not shown).

**Table 2.** Summary estimates of sensitivity and specificity with 95% confidence intervals

| Parameter | IgM | IgG | Total Ab |
| --- | --- | --- | --- |
| Sensitivity<br>Summary estimate (95%CI) | 0.82 (0.75-0.88); I<sup>2</sup>=72% | 0.85 (0.73-0.93); I<sup>2</sup>=88% | 0.85 (0.74-0.94); I<sup>2</sup>=79% |
| Specificity<br>Summary estimate (95%CI) | 0.98 (0.92-1.00); I<sup>2</sup>=92% | 0.99 (0.98-1.00); I<sup>2</sup>=13% | 0.99 (0.98-1.00); I<sup>2</sup>=74% |

## Discussion

While some SARS-CoV-2 serological tests reported an excellent ability to discriminate between seroconverted and non-seroconverted individuals, others showed diagnostic accuracy far from optimal. In particular, the pooled sensitivity was unsatisfactory (82–85%), as a substantial fraction (one sixth on average) of seroconverted individuals would be incorrectly classified as non-seroconverted. Specificity was generally very high (≥98%), yet this may not suffice to guarantee satisfactory real-world performance in areas with a very low prevalence of infected individuals. A specificity just less than perfect (99%) would in fact produce a PPV ranging between 76% and 88% when combined with a true prevalence equal to 5%, meaning that around one fifth of those labelled as seroconverted would in reality be false positives. According to WHO, 2–3% of the global population may have been infected by the end of the first epidemic wave [13], thus the PPV in most areas could indeed be much lower than in our simulations. Further reasons of concern lie in the low number of subjects on which some estimates are based, the fact that some of the included studies have not been peer-reviewed yet, the variability in terms of the gold standard used to define sensitivity and specificity, the possible heterogeneity of testing procedures (which should be harmonized internationally to ensure comparability), and, above all, the uncertainty as to whether positivity to the test means that effective protection against re-infection has been established [14–15]. Moreover, issues of cost, speed, and availability should also be taken into account when planning large seroprevalence surveys, as well as the medical and non-medical costs of diagnostic errors. While the currently available serological tests can be used for research purposes, our data suggest that their use for large-scale prevalence surveys (or to grant “immunity passports”) are currently only justified (and only if showing very high diagnostic accuracy) in hard-hit regions, while they should be used with caution elsewhere. Finally, SARS-CoV-2 serological tests are being developed at a fast pace, and these conclusions may need revision in the coming months, also depending on the further spread of the pandemic.

## Data Availability

This is a systematic review and meta-analysis, all the data is publically available to researchers

## Acknowledgments

There was no funding for this manuscript.

